# Impacts of the COVID-19 pandemic on deprivation-level differences in cardiovascular hospitalisations: A comparison of England and Denmark using the OpenSAFELY platform and National Registry Data

**DOI:** 10.1101/2024.05.09.24307105

**Authors:** Ruth E Costello, Alasdair D Henderson, John Tazare, Lars Pedersen, Henrik Toft Sorensen, Jan P Vandenbroucke, Kathryn E Mansfield, Viyaasan Mahalingasivam, Bang Zheng, Helena Carreira, Patrick Bidulka, Dominik Piehlmaier, Angel YS Wong, Charlotte Warren-Gash, Joseph F Hayes, Jennifer K Quint, Srinivasa Vittal Katikireddi, Brian MacKenna, Amir Mehrkar, Sebastian Bacon, Ben Goldacre, Laurie Tomlinson, Sinéad M Langan, Rohini Mathur, The LH&W NCS (or CONVALESCENCE) Collaborative and the OpenSAFELYcollaborative

## Abstract

**Objectives:** To examine the impact of the pandemic on deprivation-related inequalities in hospitalisations for CVD conditions in Denmark and England between March 2018 and December 2021.

**Design:** A series of monthly cross-sectional studies separately in England and Denmark. **Setting:** With the approval of NHS England, we used English primary care electronic health records, linked to secondary care and death registry data through the OpenSAFELY platform, and nationwide Danish health registry data.

**Participants:** Adults aged 18 and over, without missing age, sex or deprivation information were included. On 1st March 2020, 16,234,700 people in England, and 4,491,336 people in Denmark met the inclusion criteria.

**Primary and secondary outcome measures:** Hospital admissions with the primary reason myocardial infarction (MI), ischaemic or haemorrhagic stroke, heart failure, and venous thromboembolism (VTE).

**Results:** We saw deprivation gradients in monthly CVD hospitalisations in both countries, with differences more pronounced in Denmark. Based on pre-pandemic trends, in England, there were an estimated 2608 fewer admissions than expected for heart failure in the most deprived quintile during the pandemic, compared to an estimated 979 fewer admissions in the least deprived quintile. In Denmark, there were an estimated 1013 fewer admissions than expected over the pandemic for MI in the most deprived quintile compared to 619 in the least deprived quintile. Similar trends were seen for stroke and VTE, though absolute numbers were smaller.

**Conclusions:** Overall, we did not find that the pandemic substantially worsened pre-existing deprivation-related differences in CVD hospitalisations, though there were exceptions in both countries.

**Strengths and limitations:**

- This was one of the largest studies of the impact of the pandemic on deprivation inequalities, covering 20 million people in two countries (England and Denmark).
- Followed-up was until the end of 2021, which is longer than most previous studies.
- We compared the impact in two countries that have free at the point of use healthcare, but different responses to the pandemic.
- The measures of deprivation were different in the two countries, with the measure in England (Index of Multiple Deprivation 2019) capturing more aspects of deprivation compared to the Danish measure (income) which may have resulted in misclassification.
- Our results are descriptive so do not provide insight into the causes of observed differences.

## Introduction

Cardiovascular disease (CVD) is the leading cause of death worldwide, accounting for one in four deaths in the UK (1). CVD is known to be associated with important ethnic and socioeconomic health inequalities. Individuals living in deprived areas are more likely to have CVD and have a higher risk of dying from CVD compared with those living in the least deprived areas (2–4).

While the direct effects of the COVID-19 pandemic have been found to disproportionately affect older people, global majority ethnic groups, and deprived populations, inequalities in the indirect effects of the pandemic have yet to be fully explored (5–8). Diversion of healthcare resources to pandemic management has negatively affected non-COVID-related healthcare provision, including prevention activities, potentially creating or worsening physical and mental health (9). The negative impacts of the pandemic have been compounded by the rising cost-of-living crisis which may have further widened socioeconomic inequalities (10). One systematic review examining the impact of the COVID-19 pandemic on CVD related care (11) highlighted reduced and delayed CVD-related hospital admissions, except for cardiac arrests, and increased CVD mortality. However, a Swiss study of deprivation and CVD found that there were no changes in the relative patterning of inequalities resulting from the pandemic (12).

The UK experienced one of the worst COVID-19 outbreaks in Europe. Several Scandinavian countries have experienced better COVID-19 outcomes and faster healthcare system recovery (13). Confirmed COVID-19 deaths were higher in the UK compared to Denmark and the stringency of COVID-19 measures such as school closures, workplace closures and travel bans were higher in the UK compared to Denmark (14) supplementary materials. We do not know which aspects of the pandemic have been driving non-COVID health consequences. Comparing countries with different pandemic curves, where different measures were taken at different times may offer insights into which factors, if any, drive between-country differences, potentially informing policy for future infectious disease outbreaks.

We aimed to examine the impact of the pandemic on deprivation-related inequalities in hospitalisations for CVD conditions in Denmark and England between March 2018 and December 2021.

## Methods

Using electronic health record and registry data we conducted a series of monthly cross-sectional studies separately in England and Denmark. The cohorts for each country were defined using comparable inclusion criteria, exposure and outcome definitions, and the same statistical analysis techniques were applied (Table 1).

**Table 1:** Summary of English and Danish study designs.

|  | England | Denmark |
| --- | --- | --- |
| Inclusion criteria | Adults aged 18 and over, registered with a GP for at least 3 months prior to study entry. | Adults aged 18 and over, recorded and alive at cohort entry according to the Civil Registration System. |
| Exclusion criteria | Missing age, sex, or patient level IMD, household size >15 or household size missing. | Missing age, sex, or income. |
| Denominator population entry point | Latest of: meeting inclusion criteria or 1st March 2018. | Latest of: meeting inclusion criteria or 1st March 2018. |
| Denominator population exit point | Earliest of death, deregistering with their GP or end of study period. | Earliest of death, emigration according to the Civil Registration System or end of study period. |
| Exposure |  |  |
| Deprivation measurement | Deprivation quintiles based on IMD in the month of interest. | Deprivation quintiles based on household income in 2020. |
| Outcomes |  |  |
| Hospital admissions | Hospital admissions with ICD-10 code for heart failure, MI, stroke or VTE, as the primary reason for admission (this refers to primary reason for spell in hospital) | Hospital admissions with ICD-10 code for heart failure, MI, stroke or VTE as the primary reason for admission |

Abbreviations: GP = general practitioner, IMD = index of multiple deprivation, ICD-10 = International Classification of Diseases: Version 2010, MI = myocardial infarction, VTE = venous thromboembolism.

### Data Sources

In England, we used: 1) primary care records managed by the general practice software provider TPP; 2) Office for National Statistics (ONS) death register data; and 3) secondary care data from NHS Digital’s Secondary Use Service data containing information on hospitalisations. All data were linked, stored and analysed securely using the OpenSAFELY platform, https://www.opensafely.org/, as part of the NHS England OpenSAFELY COVID-19 service. The population covers 43% of the UK population and is broadly representative of the English population (15). Pseudonymised data included coded diagnoses. All code is shared openly for review and re-use under MIT open licence (https://github.com/opensafely/covid_collateral_imd). Detailed pseudonymised patient data is potentially re-identifiable and therefore not shared.

In Denmark, all residents are assigned a unique personal identification number (the CPR-number) at birth or immigration, which makes it possible to link individual information among different data sources. We used data from: 1) the Danish National Patient Registry (16), containing all inpatient discharge diagnoses from all Danish hospitals since 1977 and from emergency room and outpatient specialist clinic contacts since 1995 (Diagnoses are coded according to the International Classification of Diseases (ICD) 8 from 1977 to 1993, and to the ICD 10 thereafter); 2) the Danish Civil Registration System, including vital status and date of death for the entire Danish population; 3) socioeconomic registries maintained by Statistics Denmark, including data on family and household socioeconomics, country of origin, educational level, employment status, and income; and 4)The Danish Prescription Registry, which has recorded all redeemed drug prescriptions from community pharmacies in Denmark since 1995 (17).

### Study population

In England, the study population included adults, aged 18 and over, registered at a general practice using TPP software, with at least 3 months of continuous registration with the practice prior to study entry. In Denmark, the study population included all adults aged 18 and over registered in the Danish Civil Registration System. In both countries, we excluded people with missing age, sex or deprivation information (defined in the exposures section) as this could indicate poor data quality. In England, people were also excluded if their household size was greater than 15 to exclude people living in institutions such as care homes, who may have different hospital admission patterns. The measure of household size was a maximum of 15 in Denmark.

In both settings, the study period was 1st March 2018 and 31st December 2021. This was to give 2 years of data prior to the start of the pandemic for comparison, the study ended on 31st December 2021 as Danish data were only available up until this date. People entered the study at any time point during the study period as counts of outcomes were measured monthly. Follow-up continued until death or the end of the study period, in England to measure denominators, people would also end follow-up if they deregistered with their GP.

### Study measures

#### Exposures

The primary exposure was socio-economic deprivation measured by proxy. In England deprivation was measured using quintiles of the patient-level index of multiple deprivation (IMD) 2019 (18). IMD is a lower super output area level (comprising 400 to 1200 people) measure of relative deprivation based on a person’s postcode. The IMD score is based on indicators related to income, education, employment, health, crime, barriers to housing and services and living environment. We were unable to access an equivalent deprivation index in Denmark, so we used one aspect of deprivation; annual household income derived from the Danish Income Statistics Registry and divided into quintiles by year of age, due to the variations in income by age (see supplemental materials for details) (19).

Differences in outcomes by deprivation quintile were compared before and after the start of the pandemic restrictions. In England pandemic restrictions were imposed on 23rd March 2020 (20), equivalent restrictions were imposed in Denmark on 11th March 2020 (21). Since behaviours were likely to have changed prior to these dates we used 1st March 2020 as the cut-off for both countries, with time before this date referred to as the pre-pandemic period.

#### Outcomes

In both countries, we identified CVD-related hospital admissions, based on recorded International Classification of Diseases Version 10 (ICD-10) codes for myocardial infarction (MI), ischaemic or haemorrhagic stroke, heart failure, and venous thromboembolism (VTE) assigned as the primary reason for admission.

#### Demographic and clinical characteristics

Demographic characteristics were identified at 3 time-points to describe the cohorts, these included age categorised into 20-year age bands, sex and, in England only, rural-urban classification. In England, comorbidities were identified from primary care records. People with a SNOMED CT code for type 1 or type 2 diabetes mellitus on or before each time-point were considered to have diabetes. People with a SNOMED CT code for asthma in the 3 years prior to each time-point were considered to have asthma. People aged 40 years or over with a SNOMED CT code for COPD were considered to have COPD. In Denmark, where clinical diagnosis data from primary care are not available, definitions for diabetes, asthma, and COPD were based on hospital discharge diagnoses, as well as primary-care prescribing data from the Prescription Registry.

### Statistical analysis

The characteristics of each cohort, overall and by deprivation quintile, were described on 1st March 2019, 2020 and 2021. On the first day of each month of follow-up (from March 2018 to December 2022, inclusive), the inclusion criteria were assessed and the denominator adult population who met the inclusion criteria was extracted from respective national databases. Each outcome was analysed separately and individuals with outcomes were counted once each month. Individuals with records for the same outcome in multiple months were included each time.

The percentage of people experiencing each outcome was calculated for every study month. We plotted the monthly percentage and the percentage change compared to the previous month (first derivative) by deprivation quintile. To estimate the absolute impact of the pandemic on each outcome, we used Poisson regression adjusted for an indicator of whether it was pre-or during the pandemic (binary), deprivation quintile, the interaction of both pandemic time and deprivation quintile. We further adjusted for population as an offset, and time as a monthly continuous variable, to estimate the average count of each outcome, by deprivation quintile, in the 22 months pre-pandemic (May 2018-February 2020) and the 22 months during the pandemic (March 2020-December 2021). We accounted for autocorrelation by including first-order lagged residuals. We used the estimated average counts from the Poisson model to generate rate differences in the numbers of each monthly outcome, stratified by deprivation quintile.

We used Python 3.9.12 for data management, and Stata 17 and R version 4.2.1 for analyses. Code for data management and analysis, as well as codelists, are archived online https://github.com/opensafely/covid_collateral_imd. All iterations of the pre-specified study protocol are archived with version control https://github.com/opensafely/covid_collateral_imd/tree/main/docs.

#### Information governance and ethical approval

In England, NHS England is the data controller of the NHS England OpenSAFELY COVID-19 Service; TPP is the data processor; all study authors using OpenSAFELY have the approval of NHS England (22). This implementation of OpenSAFELY is hosted within the TPP environment which is accredited to the ISO 27001 information security standard and is NHS IG Toolkit compliant (23). Further information can be found in the supplementary materials. This study was approved by the Health Research Authority (REC reference 20/LO/0651) and by the LSHTM Ethics Board (reference 21863).

## Results

On 1st March 2020, 16,234,700 people in England, and 4,491,336 people in Denmark, met the inclusion criteria. The characteristics of the study populations were similar, though there were differences in the recorded prevalence of comorbidities. There was a higher recorded prevalence of diabetes in England (England: 7.9% versus Denmark: 6.5%), and a higher recorded prevalence of asthma and COPD in Denmark (Table 2). Study population characteristics were similar in 2019 and 2021 (supplementary materials).

**Table 2:** Characteristics of English and Danish study populations as of 1st March 2020.

| Characteristic |  | England*<br>N=16,439,645<br>n (%) | Denmark<br>N=4,491,336<br>n (%) |
| --- | --- | --- | --- |
| Age category | 18 - 40 years | 5,908,145<br>(35.9) | 1,600,989<br>(35.7) |
|  | 41 - 60 years | 5,390,450<br>(32.8) | 1,543,305<br>(34.4) |
|  | 61 - 80 years | 4,094,795<br>(24.9) | 1,145,112<br>(25.5) |
|  | >80 years | 1,046,255 (6.4) | 201,930 (4.5) |
| Sex | Female | 8,330,335<br>(50.7) | 2,209,312<br>(49.1) |
|  | Male | 8,109,310<br>(49.3) | 2,282,024<br>(50.8) |
| Deprivation<sup>\$</sup> | 1 (Most deprived) | 3,230,685<br>(19.7) | 864,398 (19.3) |
|  | 2 | 3,297,365<br>(20.1) | 903,277 (20.1) |
|  | 3 | 3,570,705<br>(21.7) | 906,819 (20.2) |
|  | 4 | 3,324,625<br>(20.2) | 908,303 (20.2) |
|  | 5 (Least deprived) | 3,016,270<br>(18.3) | 908,539 (20.2) |
| Rural-Urban | Rural | 3,513,405<br>(21.4) | - |
|  | Urban | 12,926,245<br>(78.6) | - |
| Diabetes mellitus<sup>&</sup> |  | 1,309,600 (7.9) | 292,027 (6.5) |
| Asthma<sup>^</sup> |  | 1,439,760 (8.8) | 619,136 (13.8) |
| COPD<sup>%</sup> |  | 533,645 (3.2) | 417,649 (9.3) |

\*England data is rounded to the nearest 5.
<sup>\$</sup> Deprivation measured by Index of Multiple Deprivation in England and income in Denmark.
<sup>^</sup> Asthma definition: England: asthma code in primary care record in the 3 years prior to study entry, Denmark: hospital diagnosis code or asthma medication prescribing
<sup>&</sup> Diabetes definition: Type 1 or type 2 diabetes mellitus code in primary care record prior to study entry, Denmark: hospital diagnosis code or diabetes medication prescribing
<sup>%</sup> COPD definition: England: age >40 with COPD code in primary care record prior to study entry, Denmark: hospital diagnosis code, or COPD medication prescribing.

When stratified by deprivation quintile, in England people in the most deprived quintile were younger with 44% aged 18-40 years old versus 28.8% of the least deprived quintile. In Denmark, age was taken into account in the deprivation quintiles, therefore age distributions were similar across deprivation quintiles. In both countries, COPD and diabetes were more prevalent in the most deprived quintile (COPD: England: most deprived: 4.6% versus least deprived: 2.3%, Denmark: most deprived: 11.5% versus least deprived: 7.6%, diabetes:

England: most deprived: 9.5% versus least deprived: 6.7%, Denmark: most deprived: 8.8%, least deprived: 4.3%).

### Hospital admissions overall

In both countries, there were similar proportions of the population admitted to hospital for each CVD outcome, although patterns by deprivation level differed between countries.

In England, across all outcomes, differences by deprivation level were small, although people in the most deprived quintile had the highest percentage of admissions for all outcomes. Across all outcomes, we observed a drop in admissions at the start of the pandemic and then a recovery to at least pre-pandemic levels by August 2020. This pattern did not vary by deprivation level. The largest decline in admissions was for heart failure. (Figure 1A and supplementary materials).

In Denmark, variation by deprivation quintile was more pronounced than in England for all outcomes. Overall, individuals in the most deprivation quintile had the highest proportion of admissions with admissions decreasing with decreasing deprivation. The biggest deprivation-related differences were seen for heart failure. The drop in admissions in March 2020 was greatest for individuals in the most deprived quintile, with smaller drops seen in the less deprived quintiles. (Figure 1B and supplementary materials).

**Figure 1A:**
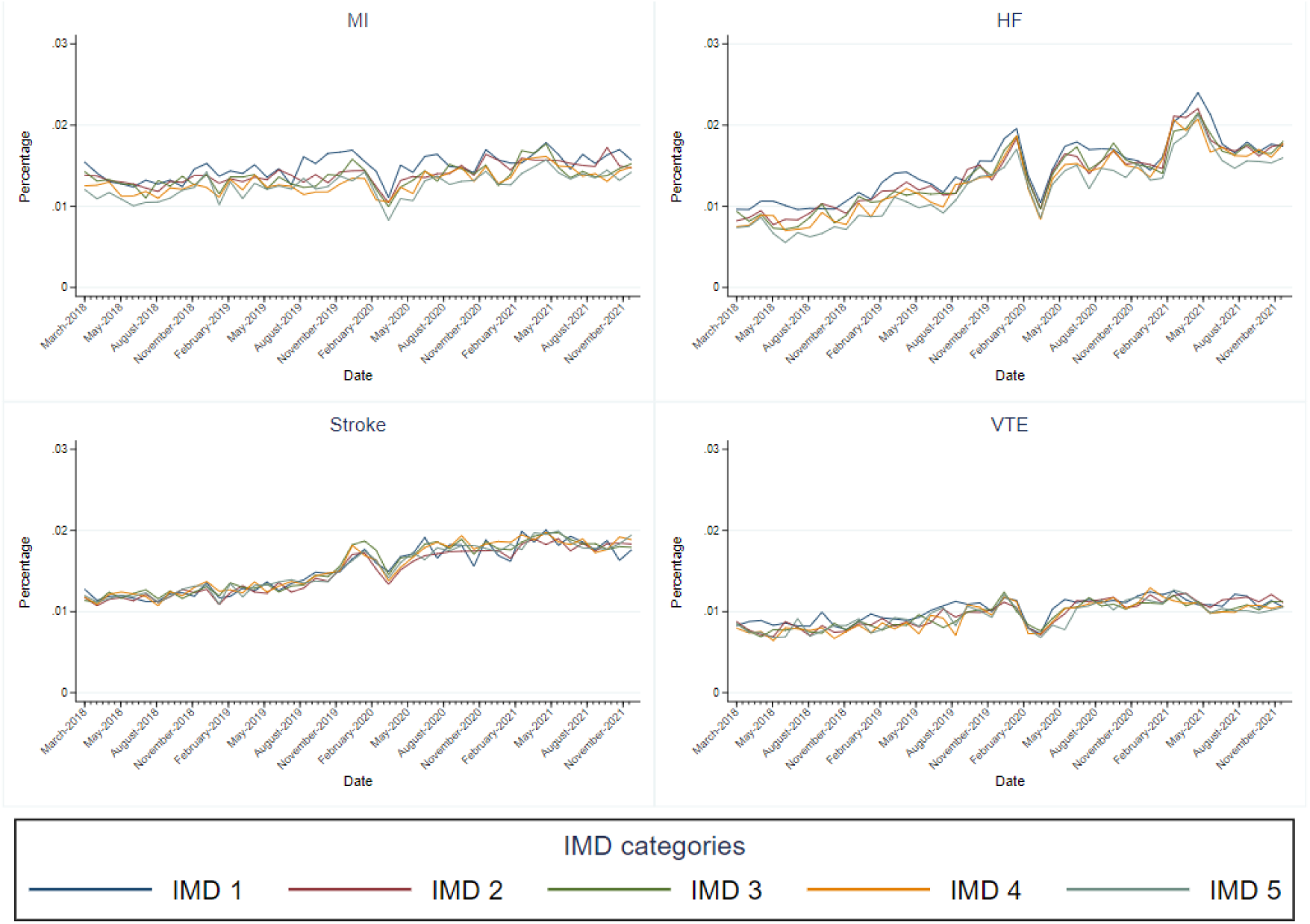
Monthly percentage of population with hospital admissions for a) myocardial infarction, b) stroke, c) heart failure, d) venous thromboembolism, by deprivation quintile, in England

**Figure 1B:**
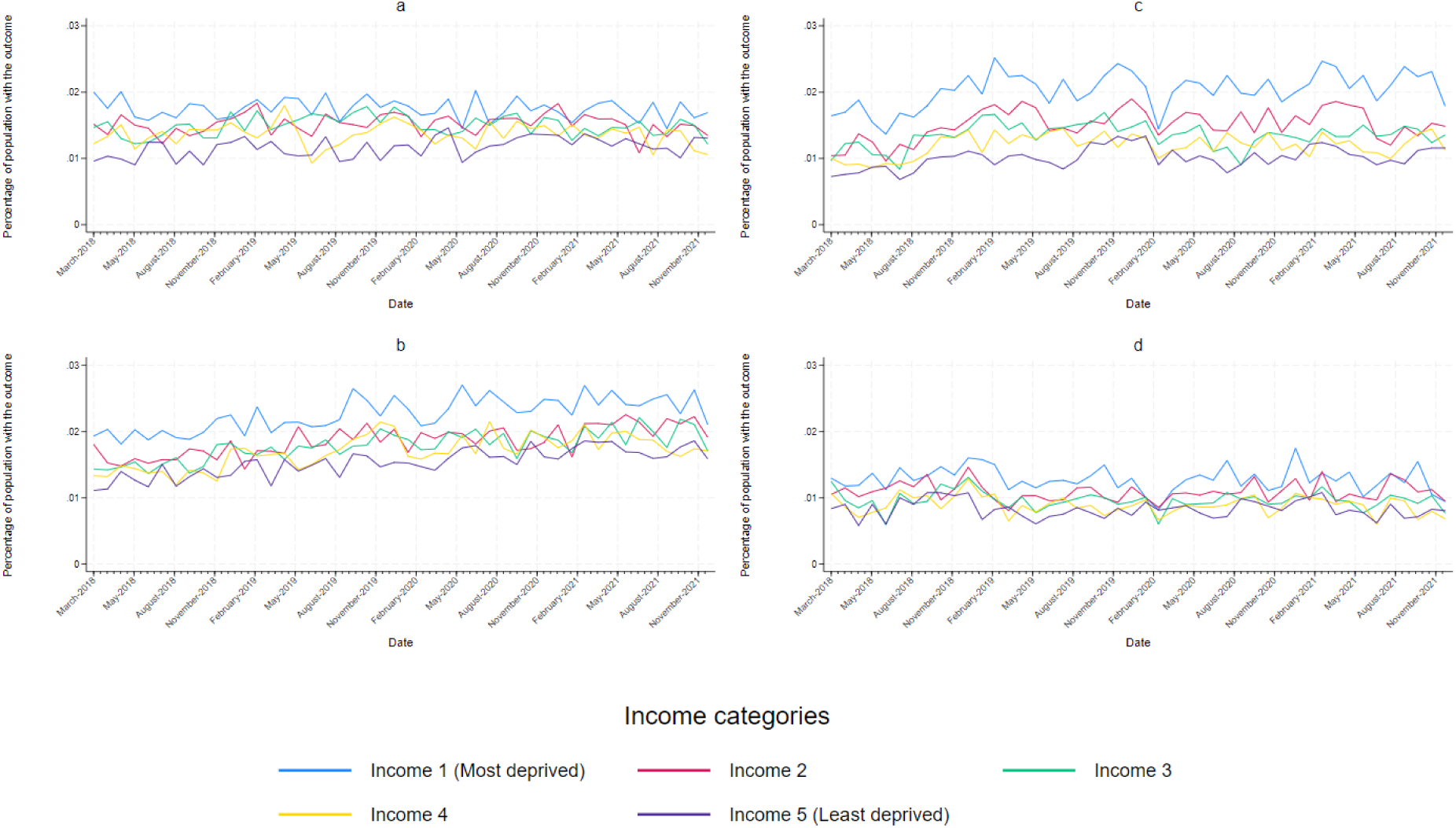
Monthly percentage of population with hospital admissions for a) myocardial infarction, b) stroke, c) heart failure, d) venous thromboembolism, by deprivation quintile, in Denmark

### Hospitalisations during the pandemic

Poisson regression models indicated that, within deprivation quintiles, the number of admissions during the pandemic (1st March 2020 to 31st December 2021) was lower than expected, and that there were small deprivation gradients in both England and Denmark.

#### England

In England, admissions for heart failure, MI and VTE were lower than expected, with the gap between observed and expected largest for people in the most deprived quintile and smallest for those in the least deprived quintile. For heart failure admissions, the gap between observed and expected admissions was largest for individuals in the most deprived quintile and narrowed with decreasing deprivation. For people living in areas classified in the most deprived quintile, heart failure admissions were 17.8% lower than expected, which in absolute terms translated to an estimated 2608 fewer admissions between March 1st 2020 and December 31st 2021. In the least deprived quintile heart failure admissions were 9% lower than expected, translating to an estimated 979 fewer admissions between March 1st 2020 and December 31st 2021. For MI, variation by deprivation level followed a similar pattern, although differences were smaller. For VTE there were estimated to be fewer admissions than expected, though there was little variation by deprivation quintile. For stroke there were slightly more admissions than expected, also with little variation by deprivation quintile (Figure 2A and Table 3).

**Table 3A:**
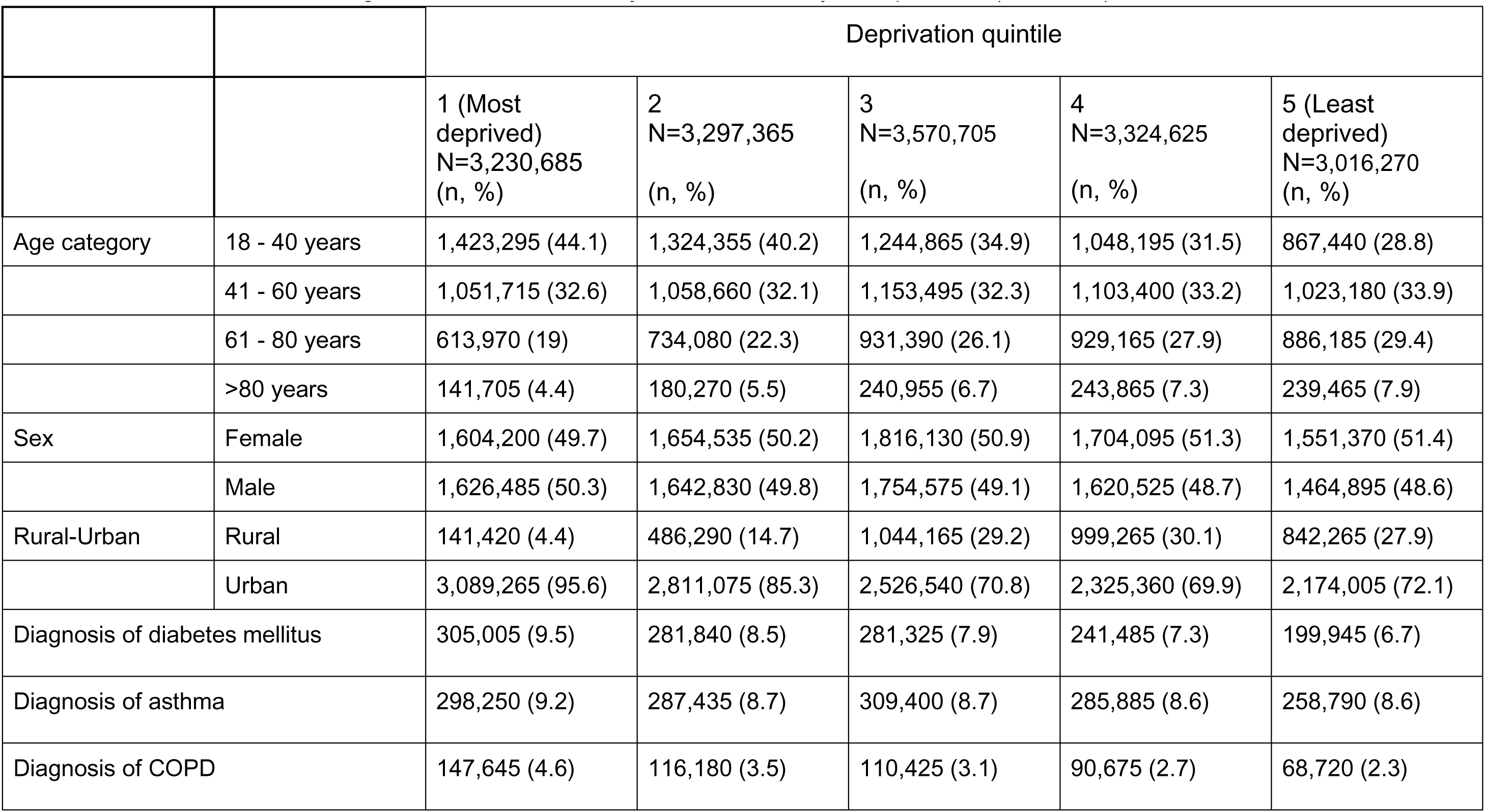
Characteristics of the English cohort on 1st January 2020, stratified by IMD quintile, n (column %)

**Table 3B:** Characteristics of the Danish cohort on 1st January 2020, stratified by deprivation quintile.

|  |  | Deprivation quintile |  |  |  |  |
| --- | --- | --- | --- | --- | --- | --- |
|  |  | 1 (Most deprived)<br>N= 864,398<br>(n, %) | 2<br>N= 903,377<br>(n, %) | 3<br>N= 906,819<br>(n, %) | 4<br>N= 908,303<br>(n, %) | 5 (Least deprived)<br>N= 908,539<br>(n, %) |
| Age category | 18 - 40 years | 292,006 (33.8) | 324,187 (35.9) | 327,324 (36.1) | 328,475 (36.2) | 328,997 (36.2) |
|  | 41 - 60 years | 303,906 (35.2) | 309,437 (34.3) | 309,854 (34.2) | 310,154 (34.1) | 309,954 (34.1) |
|  | 61 - 80 years | 228,127 (26.4) | 229,234 (25.4) | 229,232 (25.3) | 229,289 (25.2) | 229,230 (25.2) |
|  | >80 years | 40,359 (4.7) | 40,419 (4.5) | 40,409 (4.5) | 40,385 (4.4) | 40,358 (4.4) |
| Sex | Female | 448,832 (51.9) | 495,897 (54.9) | 454,872 (50.2) | 444,284 (48.9) | 438,139 (48.2) |
|  | Male | 415,566 (48.1) | 407,380 (45.1) | 451,947 (49.8) | 464,019 (51.1) | 470,400 (51.8) |
| Rural-Urban | Rural | - | - | - | - | - |
|  | Urban | - | - | - | - | - |
| Diagnosis of diabetes mellitus |  | 76,420 (8.8) | 67,905 (7.5) | 59,439 (6.6) | 49,510 (5.5) | 38,753 (4.3) |
| Diagnosis of asthma |  | 113,871 (13.2) | 126,173 (14.0) | 128,184 (14.1) | 127,351 (14.0) | 123,557 (13.6) |
| Diagnosis of COPD |  | 99,289 (11.5) | 91,384 (10.1) | 82,567 (9.1) | 75,052 (8.3) | 69,357 (7.6) |

#### Denmark

In Denmark, admissions for MI were lower than expected. As a proportion of the number of expected admissions, the gap between observed and expected admissions over the pandemic period was largest for people in the least deprived quintile, where admissions were 24% lower than expected compared to the most deprived quintile, where admissions were 22% lower than expected. However, in absolute terms, differences were greatest in the most deprived quintile with 1013 fewer admissions during the pandemic compared to 619 fewer admissions in the least deprived quintile. For all other outcomes admissions during the pandemic were similar to pre-pandemic levels, with little variation by deprivation level (Figure 2B and Table 4).

**Figure 2A:**
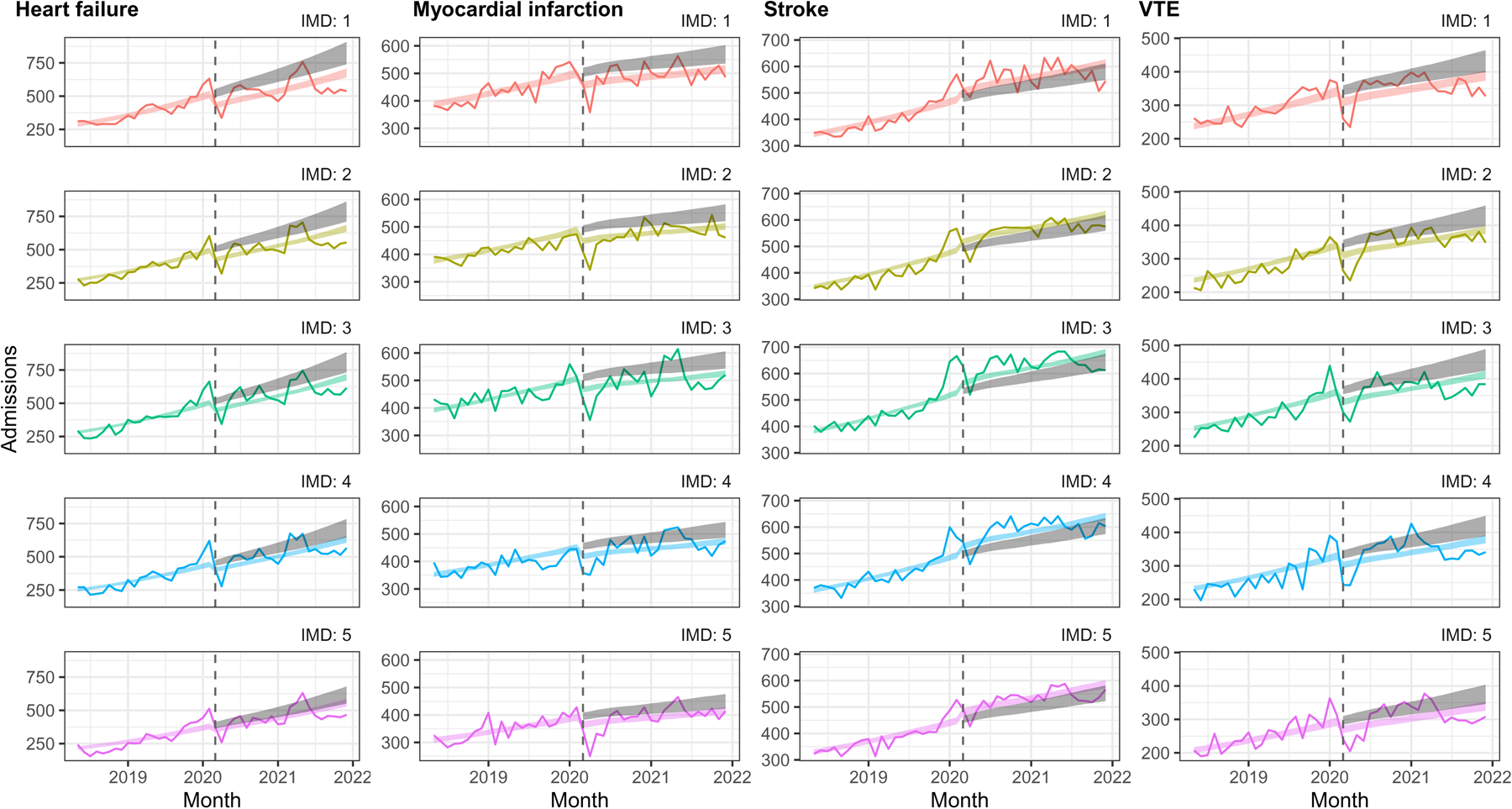
Interrupted time-series analysis of changes in hospital admissions in England before the pandemic (May 2018-February 2020) compared to during the pandemic (March 2020 to December 2021), by deprivation quintile. Coloured lines indicate the estimated number of admissions per month with COVID-19 restrictions, grey lines indicate the estimated number of admissions per month without COVID-19 restrictions.

**Figure 2B:**
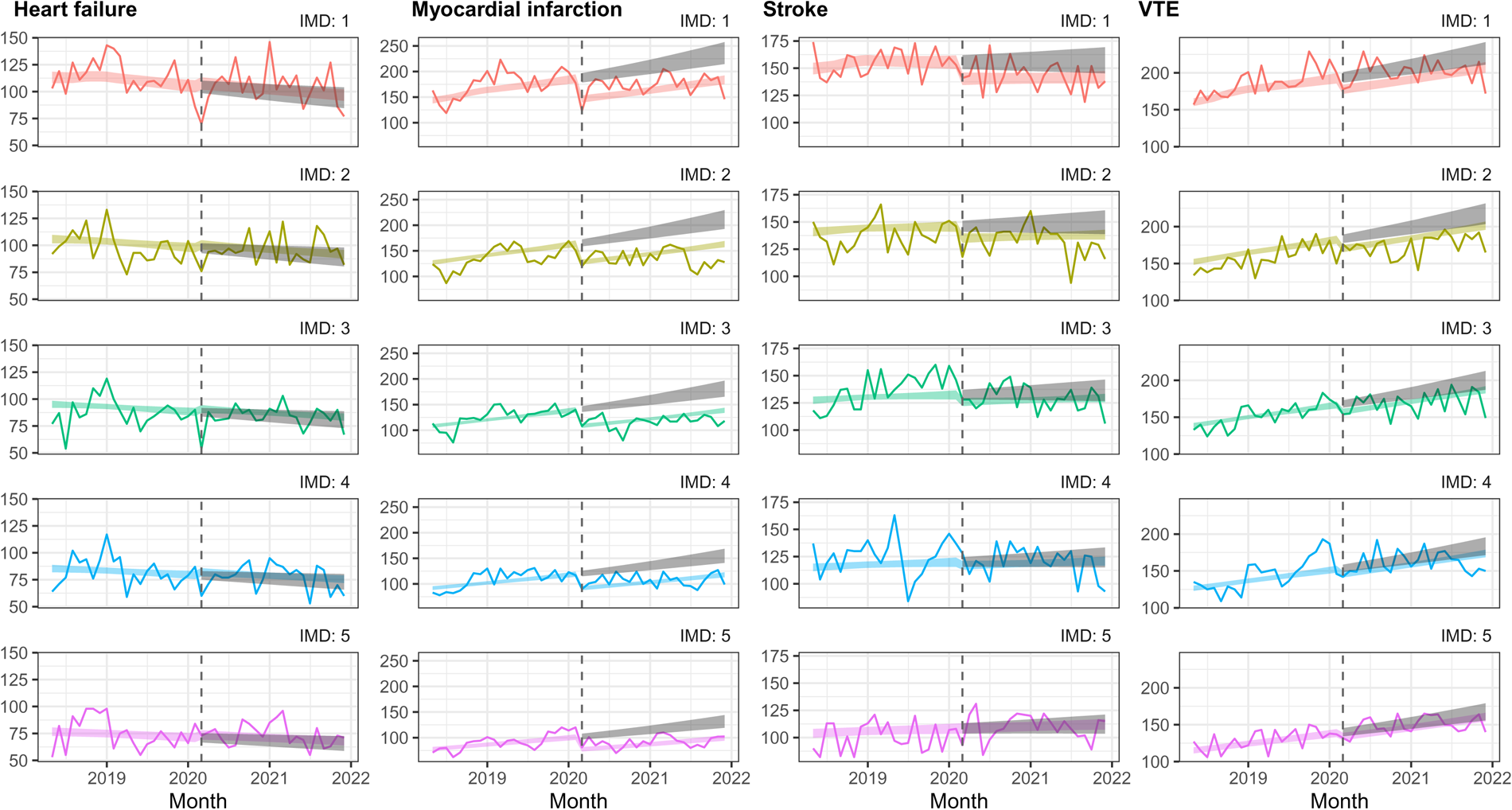
Interrupted time-series analysis of changes in hospital admissions in Denmark before the pandemic (May 2018-February 2020) compared to during the pandemic (March 2020 to December 2021), by deprivation quintile. Coloured lines indicate the estimated number of admissions per month with COVID-19 restrictions, grey lines indicate the estimated number of admissions per month without COVID-19 restrictions.

## Discussion

In this observational study set in England and Denmark, we found that deprivation-level differences in cardiovascular hospitalisations were not exacerbated by the pandemic, with a few exceptions. In England, overall, there were fewer heart failure admissions during the pandemic than expected, and reductions increased with increasing deprivation. In Denmark there were fewer stroke and VTE admissions than expected during the pandemic in the most deprived quintile. In England, overall cardiovascular admissions increased over time whereas in Denmark admissions remained stable.

In England, we observed a deprivation gradient across our outcomes, which was comparable to that observed for other health outcomes (3). However, differences by deprivation level were substantially more marked in Denmark. This could be due to the different measures of deprivation used. In Denmark, we used household-level income, while in England, we used IMD (a small area level measure based on the average deprivation level of an area, assessed across a range of seven domains including income). IMD’s sensitivity and specificity to income deprivation is low (24), some people’s deprivation levels could have been misclassified. Assuming such misclassification was not differential, this could bias any differences towards the null, which could explain the smaller differences between deprivation levels in England compared to Denmark.

Compared to the expected admissions, reductions in actual admissions between the pre-pandemic and pandemic periods were greater in England compared to Denmark, which generally experienced little change. This is consistent with other studies of CVD admissions and specifically for non-ST-elevation acute coronary syndromes in 2020 (7,25) Our study updates these findings to demonstrate that this pattern continued into 2021. The restrictions in Denmark were less stringent than in England (supplementary materials) and there were fewer COVID-19 deaths (26). This may have meant cardiology services in hospitals remained similar during the pandemic as the health service may not have been so overwhelmed, whereas in England there was extreme disruption to primary care and secondary care cardiology services, which would affect preventative care (27) and health seeking for acute CVD events. In addition some heart failure services moved into the community in England, which may have resulted in fewer hospital admissions (28).

Although studies have investigated the impact of the pandemic on cardiovascular admissions (7,25), only a few studies have specifically investigated whether the pandemic impacted cardiovascular admissions by deprivation level (29–31). Two studies, in the USA and Catalonia, compared socioeconomic differences in heart failure admissions between 2019 and 2020 found that the impact of the pandemic was similar across income groups (26,29). These results are similar to our findings from Denmark where the impact of the pandemic was similar across deprivation groups, in contrast to England where the reduction in heart failure admissions during the pandemic was larger in the most deprived. One study set in the USA found that the impact of the pandemic on stroke admissions was similar across income groups (30). This was consistent with our findings in England, whereas in Denmark there were slightly fewer admissions during the pandemic in the most deprived group but differences were small. As these studies are set in different countries, there could be many reasons for the observed differences in admissions.

### Strengths and limitations

Our study was large, encompassing 20 million people across two countries. Our study period was until the end of 2021, longer than most previous studies (which largely ended in 2020) (29–31), allowing us to describe the longer-term impacts of the pandemic. Our study design allowed us to compare the impact in two countries that both have a free-at-the-point-of-use health service, but different responses to the pandemic. However, an important limitation was that our measures of deprivation were different in the two countries, with the measure in

England capturing more aspects of deprivation than the Danish measure, resulting in potential misclassification. Another limitation was that some information was not available in both countries, thus we could not examine cardiovascular mortality or ethnicity as this was unavailable in Denmark. Finally, since our results are descriptive, they do not provide insight into the causes of any observed differences.

## Conclusions

During the pandemic we did not observe a worsening of socioeconomic gradient on cardiovascular admissions in England and Denmark. There were some exceptions, most notably greater reductions in heart failure admissions in the most deprived groups in England. While it is positive that the pandemic has not worsened socioeconomic differences in cardiovascular admissions, further work is needed to understand the reasons for the differences seen in heart failure admissions in England.

## Data Availability

Access to the underlying identifiable and potentially re-identifiable pseudonymised electronic health record data is tightly governed by various legislative and regulatory frameworks, and restricted by best practice. The data in the NHS England OpenSAFELY COVID-19 service is drawn from General Practice data across England TPP is the data processor.
TPP developers initiate an automated process to create pseudonymised records in the core OpenSAFELY database, which are copies of key structured data tables in the identifiable records. These pseudonymised records are linked onto key external data resources that have also been pseudonymised via SHA-512 one-way hashing of NHS numbers using a shared salt. University of Oxford, Bennett Institute for Applied Data Science developers and PIs, who hold contracts with NHS England, have access to the OpenSAFELY pseudonymised data tables to develop the OpenSAFELY tools.
These tools in turn enable researchers with OpenSAFELY data access agreements to write and execute code for data management and data analysis without direct access to the underlying raw pseudonymised patient data, and to review the outputs of this code. All code for the full data management pipeline, from raw data to completed results for this analysis, and for the OpenSAFELY platform as a whole is available for review at github.com/OpenSAFELY.

## Acknowledgements

Thanks to Dr Edward Parker for his help with output checking. This work uses data provided by patients and collected by the UK NHS as part of their care and support. We are very grateful for all the support received from the TPP Technical Operations team throughout this work, and for generous assistance from the information governance and database teams at NHS England and the NHS England Transformation Directorate. We have a publicly available website https://opensafely.org/ where we invite individuals to contact us regarding this study or the broader OpenSAFELY project.

## Contributors

REC, JT, ADH, LP, SL & RM were involved in the development of the study. REC, ADH, JT, LP, HTS, VM, BZ, BM, SB and RM had access to the data. REC, ADH, JT, VM, BZ and RM verified the underlying data. REC, ADH and JT were responsible for data management and statistical analysis. REC and RM wrote the first draft of the manuscript. All authors contributed to and approved the final manuscript, and accept responsibility to submit for publication.

## Data sharing

Access to the underlying identifiable and potentially re-identifiable pseudonymised electronic health record data is tightly governed by various legislative and regulatory frameworks, and restricted by best practice. The data in the NHS England OpenSAFELY COVID-19 service is drawn from General Practice data across England TPP is the data processor.

TPP developers initiate an automated process to create pseudonymised records in the core OpenSAFELY database, which are copies of key structured data tables in the identifiable records. These pseudonymised records are linked onto key external data resources that have also been pseudonymised via SHA-512 one-way hashing of NHS numbers using a shared salt. University of Oxford, Bennett Institute for Applied Data Science developers and PIs, who hold contracts with NHS England, have access to the OpenSAFELY pseudonymised data tables to develop the OpenSAFELY tools.

These tools in turn enable researchers with OpenSAFELY data access agreements to write and execute code for data management and data analysis without direct access to the underlying raw pseudonymised patient data, and to review the outputs of this code. All code for the full data management pipeline — from raw data to completed results for this analysis — and for the OpenSAFELY platform as a whole is available for review at github.com/OpenSAFELY.

